# Asymptomatic carriage rates and case-fatality of SARS-CoV-2 infection in residents and staff in Irish nursing homes

**DOI:** 10.1101/2020.06.11.20128199

**Authors:** SP Kennelly, AH Dyer, R Martin, SM Kennelly, A Martin, D O’Neill, A Fallon

**Affiliations:** Department of Age-Related Healthcare, Tallaght University Hospital, Dublin 24, Ireland; Department of Medicine for the Elderly, Connolly Hospital, Blanchardstown, Dublin 15, Ireland; Department of Geriatric and Stroke Medicine, Beaumont Hospital, Dublin 9, Ireland; Department of Age-Related Heathcare, Tallaght University Hospital, Dublin 24, Ireland

## Abstract

**Background:** SARS-CoV-2 has disproportionately affected nursing home (NH) residents. In Ireland, the first NH case of COVID-19 occurred on 16/03/2020. A national point-prevalence testing program of all NH residents and staff took place from 18/04/2020-05/05/2020.

**Aims:** To examine characteristics of NHs across three Community Health Organisations (CHOs) in Ireland, proportions with COVID-19 outbreaks, staff and resident, symptom-profile and resident case-fatality.

**Methods:** Forty-five NHs surveyed across three CHOs requesting details on occupancy, size, COVID-19 outbreak, timing of outbreak, total symptomatic/asymptomatic cases, and outcomes for residents from 29/02/2020-22/05/2020.

**Results:** Surveys were returned from (62.2%, 28/45) of NHs (2043 residents, 2303 beds). Three-quarters (21/28) had COVID-19 outbreaks (1741 residents, 1972 beds). Median time from first case of COVID-19 in Ireland to first case in these NHs was 27.0 days. Resident COVID-19 incidence was (43.9%, 764/1741): laboratory-confirmed (40.1%, 710/1741) with (27.2%, 193/710 asymptomatic), and clinically-suspected (3.1%, 54/1741). Resident case-fatality was (27.6%, 211/764) for combined laboratory-confirmed/clinically-suspected COVID-19. Similar proportions of residents in NH with an “early” outbreak (<28days) versus a later outbreak developed confirmed/suspected COVID-19. A lower proportion of residents in NHs with “early” outbreaks had recovered compared to those with “late” outbreaks (37.4% vs 61.7%; χ2=56.9, p<0.001). Among 675 NH staff across twenty-four sites who had confirmed/suspected COVID-19 (23.6%, 159/675) were asymptomatic. There was a significant correlation between the proportion of staff with symptomatic COVID-19 and resident numbers with confirmed/suspected COVID-19 (Spearman’s rho=0.81, p<0.001).

**Conclusion:** This study demonstrates COVID-19 impact on NH residents and staff. High infection rates lead to challenges in care provision.

## Background

SARS-CoV-2 infection and the related illness COVID-19 has disproportionately affected nursing home (NH) residents since its emergence in late 2019 [1]. NH residents are among the frailest in society, with multiple comorbidities and high levels of care needs. NH facilities vary in size, staffing and governance structure, and their integration with the wider health system locally, nationally, and internationally [2]. Policies on testing and reporting of COVID-19 cases in NHs differ between countries but deaths in care home residents have accounted for 24-82% of all COVID-19-related deaths [1]. Guidance published specifically for long term care facilities on infection prevention and management in the context of COVID-19 [3,4] can be challenging to implement due to variability in facilities and governance structures.

In Ireland, there is a “mixed market” of NH care provision. Public NHs are owned and operated by the government-funded Health Service Executive (HSE) and private NHs by individual providers or legal provider entities. All must be registered with the Health Information and Quality Authority (HIQA) and comply with regulations for minimum standards of care (Health Act 2007) [5]. There were 31,220 residential care beds registered across 581 NHs in Ireland in December 2018 [6]: the majority were private NHs with approximately 25,000 residents, with 121 (21%) public NHs with approximately 5,000 residents.

The first laboratory-confirmed case of COVID-19 in Ireland occurred in the community on 29/02/2020 [7]. The first clusters of COVID-19 were reported in NHs on 16/03/2020. The incidence of COVID-19 in long-term care residents in Ireland was 133/1000 population on 06/05/2020. As of 20/05/2020, 258 NH clusters were reported accounting for 4,872 cases and 851 deaths. The national case-fatality figure for NH residents related to COVID-19 is 17.5%. The majority of NH clusters have occurred around Dublin/eastern Ireland, with a sizeable proportion of NH facilities outside this area unaffected by COVID-19 outbreaks to date [7]. The national case-fatality may be an underestimate, failing to capture earlier attributable NH deaths. Given emerging evidence of impact on NHs, the Irish health service directed a national point-prevalence mass COVID-19 testing program of all residents and staff in NHs from 18/04/2020-05/05/2020.

## Aims

We aimed to examine characteristics of NHs across three Irish Community Health Organisations (CHOs) in Dublin/Eastern Ireland, the proportion with outbreaks of COVID-19, symptom profile for staff and residents, and resident mortality rates.

## Methods

An information sheet and survey document was distributed to the lead Nursing and/or Medical Officer in forty-five NHs across three CHOs. This was followed by a telephone call to obtain consent and aid completion of the survey, ensuring correct interpretation of information. Data was requested for the period 29/02/2020-22/05/2020 and a deadline for returns set for one-week later. Responses were anonymised with no site, resident, or staff identifiers requested or retained.

The survey requested details on the occupancy in the NH on 29/02/2020, total number of beds, proportion of single rooms, details of COVID-19 outbreak, date of the first laboratory-confirmed COVID-19 resident and/or staff member, total number of symptomatic/asymptomatic cases, and clinical outcomes for residents. Symptoms were defined as per public health guidance (cough, fever, dyspnoea) and any “new-onset” symptoms the medical officer/general practitioner felt reflective of emerging information on atypical COVID-19 symptoms. Asymptomatic cases (without symptoms 7-days either side of test) were identified in the national point-prevalence mass testing program of residents and staff which took place from 18/04/2020-05/05/2020. Laboratory-confirmed cases were those with “detected” SARS-CoV-2 infection by Real-Time Reverse Transcription-Polymerase Chain Reaction (RT-PCR) testing on nasopharyngeal swabs. Suspected cases were based on senior medical/nursing opinion of COVID-19 symptoms, but resident was unable/unwilling to complete testing or was isolated awaiting results.

For inclusion in analysis, an outbreak was defined as ≥1 resident with laboratory-confirmed COVID-19. NHs with one to two staff members and no residents affected weren’t considered outbreaks on the basis that infection may have been acquired outside of the NH setting. An “active” outbreak was defined as being within twenty-eight days of symptom onset for most recent laboratory-confirmed case in a resident/staff member.

Data are reported as proportions (and percentages) and mean/median with standard deviation (SD)/interquartile ranges (IQR) as appropriate. Data on NH size (number of beds) and occupancy (percentage of beds) were transformed into discrete categories for between-group analysis (size: <50 beds, 51-100 beds, >100 beds; occupancy; <75%, 75-85%, 86-95% and >95%). Comparisons of proportions between groups was carried out using a Chi-square statistic. Spearman Rank correlations were used to analyse correlations between variables. All ethical standards were adhered to in the acquisition, anonymisation, and reporting of data.

## Results

Complete surveys were returned from 62.2% (28/45) of NHs with a total of 2043 residents in 2303 beds (median occupancy 96.7%, IQR: 86.0–96.6%) on 29/02/2020. During the eighty-three-day study period, 15.3% (312/2043) of residents died.

The first laboratory-confirmed case of COVID-19 in this NH series was on 16/03/2020. Median time from the first case of COVID-19 infection in Ireland to the first recorded cases in NHs (staff/residents) included in this series was 27.0 days (IQR: 22.5-40.5). Four NHs recorded no confirmed cases of COVID-19 in staff/residents. Three further NHs had ≤2 staff members and no residents with confirmed COVID-19. All other NHs (21/28, 75.0%) were being managed as active COVID-19 outbreaks at the end of the study period and are included in the study. There were 1741 residents in 1972 beds across these sites (median occupancy rate 96.7%, IQR: 86.0– 98.6%) on 29/02/2020. Of all resident deaths recorded, 96.2% (300/312) occurred in outbreak NHs with a mortality rate of 17.2% (300/1741).

### Outbreak nursing home profile

An outbreak was recorded in 75.0% (21/28) of facilities – four public and seventeen private [Table 1]. Occupancy rates at the start of the study period were 95.1% and 87.7% in public and private NHs respectively, decreasing to 75.2% in public and 73.2% in private NHs by 22/05/2020. Eight NHs (38.1%) had ≥80% single rooms in line with regulatory standards [8]. There was no association between adherence to this standard and outbreak occurrence (χ2=1.37, p=0.24).

**Table 1:** Outbreak Nursing Home profile

|  | **Public** | **Private** | **Total** |
| --- | --- | --- | --- |
|  | **N=4** | **N=17** | **N=21** |
| **Occupancy rate** |  |  |  |
| <u>Date: 29/02/2020</u> |  |  |  |
| % , n | 95.1%, 157/165 | 87.7%, 1584/1807 | 88.3%, 1741/1972 |
| (median, IQR) | (98.1%, 94.9 – 100%) | (97.6%, 85.1 – 98.6%) | (97.8%, 86.0 – 98.8%) |
| <u>Date: 22/05/2020</u> |  |  |  |
| % , n | 75.2%, 124/165 | 73.2%, 1322/1807 | 73.3%, 1446/1972 |
| (median, IQR) | (72.3%, 60.2 – 85.2%) | (80.3%, 69.4 – 86.7%) | (80.3%, 65.7 – 86.7%) |
| **NH Size (% , n)** |  |  |  |
| <50 beds, n | 4/4 | 1/17 | 5/21 |
| 50 – 100 beds | 0/4 | 7/17 | 7/21 |
| ≥100 beds | 0/4 | 9/17 | 9/21 |
| **Table 1: Outbreak Nursing Home profile** |  |  |  |

### COVID-19 in Nursing Home Residents

During the study period, 40.1% of residents in twenty-one NHs with COVID-19 outbreaks had a laboratory-confirmed diagnosis of COVID-19. Another 3.1% had clinically-suspected COVID-19 giving a total prevalence of 43.9%. Over a quarter of residents with laboratory-confirmed COVID-19 were asymptomatic [Table 2].

**Table 2:** SARS-CoV-2 infection in nursing home residents

|  | **Confirmed** | **Suspected** | **Total** |
| --- | --- | --- | --- |
| COVID-19 infection | 40.1% | 3.1% | 43.9% |
| (% total residents) | (710/1741) | (54/1741) | (764/1741). |
| Asymptomatic | 27.2% | N/A | 27.2% |
| (% confirmed COVID-19) | (193/710) |  | (193/710) |
| Case-fatality | 25.8% (183/710) | 3.7% (28/764) | 27.6% (211/764) |
| **Table 2: SARS-CoV-2 infection in nursing home residents** |  |  |  |

The case-fatality rate was 25.8% in those with laboratory-confirmed COVID-19 infection. When clinically-suspected cases were included the overall case-fatality rate was 27.6% reflecting almost three quarters (70.3%; 211/300) of all deaths recorded in these NHs. Three NHs (7.0%; 3/21) with outbreaks had no COVID-19-related deaths. Across the remaining eighteen (85.7%; 18/21), the median proportion of total residents (as per occupancy on 29/02/2020) who died as result of confirmed/suspected COVID-19 was 15.0% (range 13.1-19.4%). Non-COVID-19 mortality was similar in outbreak-affected NHs and unaffected NHs (5.1% [89/1741] vs 4.0% [12/300] χ^2^=0.71, p=0.40).

Comparisons of resident outcomes between public and private NHs are biased by low number of public NHs in the sample (5/28, 17.9%). However, there were significantly more residents with confirmed/suspected COVID-19 in the four public (106/157; 67.5%) versus seventeen private (620/1500; 41.3%) NHs experiencing an outbreak (χ2=39.6; p<0.001). Similarly, the case-fatality attributable to COVID-19 was significantly higher in public NHs (35/157; 22.3% vs 168/1500; 11.2%; χ2=16.2; p<0.001).

In eleven NHs (11/21, 52.4%) with a first confirmed COVID-19 case “early” in the course of the pandemic (before 28/03/2020), 45.4% of residents developed confirmed/suspected COVID-19 infection, compared to 42.1% in NH with a first confirmed case after this date. NHs with “early” outbreaks had a higher number of deaths expressed as a proportion of total residents but similar case-fatality rates for residents with confirmed/suspected COVID-19 as NHs with “late” outbreaks [Table 3].

**Table 3:** Residents and Staff outcomes with SARS-CoV-2 infection in nursing homes with early versus later pandemic outbreaks

|  | **Early Outbreak**<br>**(<28 days)** | **Late Outbreak**<br>**(≥ 28 days)** |  |
| --- | --- | --- | --- |
| Total residents confirmed/<br>suspected COVID-19 infection | 45.4% (425/936) | 42.1% (339/805) | $\chi^2=2.16$, $p=0.14$ |
| Confirmed/ suspected COVID-19 deaths<br>(% total residents) | 13.5% (126/936) | 10.6% (85/805) | $\chi^2=3.42$, $p=0.06$ |
| Case-fatality rate | 29.7% (126/425) | 25.1% (85/393) | $\chi^2=1.14$, $p=0.29$ |
| Residents recovered from<br>COVID-19 | 37.4% (159/425) | 61.7% (209/339) | $\chi^2=56.9$,<br>$p<0.001$ |
| Residents remaining in<br>isolation | 32.9% (140/425) | 13.3% (45/339) | $\chi^2=15.2$,<br>$p<0.001$ |
| Total staff members confirmed<br>COVID-19 (n, %) | 50.8% (357/703) | 60.7% (318/524) | $\chi^2=11.9$,<br>$p<0.001$ |
| Asymptomatic staff members<br>confirmed | 23.1% (81/350) | 24.8% (78/314) | $\chi^2 = 0.26$,<br>$p=0.61$ |

By the end of this study period 55.8% (396/710) of residents with laboratory-confirmed COVID-19 had recovered. A lower proportion had recovered in NHs with “early” outbreaks when compared to NHs with “late” outbreaks suggesting more prolonged outbreak in the “early” group. A greater proportion of residents with confirmed/suspected COVID-19 were still in isolation in the “early” vs the “late” group [Table 3]. Six NHs (6/21, 28.6%) had no residents with confirmed/suspected COVID-19 in isolation at the end of the study period; two (18.1%; 2/11) in the “early” and four (40.0%; 4/10) in the “late” outbreak group.

In ten NHs with COVID-19 where total staffing levels were recorded, the median proportion of residents with confirmed/suspected COVID-19 was 43.7% (IQR 34.6–53.4%). Two NHs had a staff/resident ratio <1, six of 1–2 and two with >2 staff/resident. In NHs with a staff/resident ratio of <1 a median of 46.7% (IQR 33.0–60.5%) of residents had confirmed/suspected COVID-19 with a median case-fatality rate of 52.0% (IQR 45.3–59%), 27.9% (IQR 17.9–38.2%) of all residents. This compared to a median proportion of confirmed/suspected COVID-19 of 48.5% (IQR 36.3–53.4%) in residents in NHs with staff/resident ratio of 1–2 and 40.3% (IQR 39.4–41.3%) in those with staff/resident ratio of >2. In NHs with a staff resident ratio of 1–2, a median of 24.8% (IQR 22.5–28.1%) of patients with confirmed/suspected COVID-19 died; a median of 10.9% (IQR 9.5–15.6%) of all residents. One of the two NHs with a staff/resident ratio of >2 had no COVID-19 related deaths during the study period.

### COVID-19 in Nursing Home Staff

During the study period, 675 NH staff across twenty-four sites (85.7%; 24/28) (21 outbreak and 3 sites with ≤2 staff affected) had confirmed/suspected COVID-19 infection. Almost a quarter (23.6%, 159/675) were asymptomatic, identified by mass point-prevalence testing. While all NHs gave details on total staff numbers with COVID-19, twelve (42.9%, 12/28) reported information relative to total staffing levels (all grades). A total of 1392 staff members worked across these twelve sites with almost a quarter (23.8%, 331/1392) reported as confirmed/suspected COVID-19. Over a quarter were asymptomatic (27.5%, 91/331). Ten of the twelve NHs (83.3%, 10/12) met criteria for an outbreak (one NH had no staff/residents with COVID-19, and another only two staff infected). In those NHs, 329/1227 (26.8%) of staff had confirmed/suspected COVID-19 infection, and over a quarter were asymptomatic (27.1%; 89/329). The median proportion of total staff with COVID-19 per outbreak site was 31.1%; (IQR 23.2–40.6%), with 19.6% (IQR: 11.8–52.3%) of those staff being asymptomatic. Across sites, there was a significant correlation between the proportion of staff with symptomatic COVID-19 and the number of residents with confirmed/suspected COVID-19 (Spearman’s rho=0.81, p<0.001). There was no significant correlation between the proportion of asymptomatic staff and number of residents with confirmed/suspected COVID-19 (Spearman’s rho=0.18, p=0.61).

Total staffing levels were reported in six NHs (6/11, 703 staff) with “early” outbreaks and four (4/10, 524 staff) with “late” outbreaks. Approximately half of staff in NH with “early” outbreaks (50.8%; 357/703,) had confirmed/suspected COVID-19 compared to 60.7% (318/524) in NHs with “late” outbreaks. Almost a quarter of staff in both “early” (23.1%; 81/350) and “late” (24.8%; 78/314) NHs were asymptomatic [Table 3].

## Discussion

This is the largest epidemiological study of NHs, residents, and staff reported in the literature to date, demonstrating the disproportionate impact of COVID-19 on this part of the health sector. Within twenty-one NH outbreak clusters in this report, 43.9% of residents and 23.8% of staff had COVID-19 over the twelve-week period (Eighty-three-days). The case-fatality rate due to suspected/confirmed COVID-19 in residents was 26.7%. The mass point-prevalence exercise and contact-tracing testing revealed asymptomatic/pre-symptomatic COVID-19 in 27.2% of residents, and 27.5% of staff. This case-fatality rate is in contrast to the overall Irish national case-fatality rate (as of 06/06/2020) of 5.6%, almost half of which are deaths attributable to NH residents. This bias in the recording of national case-fatality figures has led some commentators to suggest NH-related mortality should be examined separate to other community deaths [9].

Residents in NHs have increased comorbidities and frailty. Much of the increased mortality risk is associated with these factors. The usual annualised morality rate of residents is estimated at 31.8% or 0.087%/day [10]; in this study the mortality rate was 15.3% overall and 17.2% in outbreak NHs over eighty-three days, equating to a daily rate of 0.184% and 0.207% respectively, more than double (x2.38) the normal rate. With regards international comparisons on NH COVID-19 infection rates and case-fatality, our results are consistent with a report on eighty-nine residents from a US facility [11], with an infection rate of 64% and a case-fatality rate of 26%, and a series from 394 residents in four UK NHs with an infection rate of 40%, and a case-fatality of 26% [12]. In the UK series 43% of residents were asymptomatic, as were 4% (n=70) of staff.

This is the first report to investigate the effect of outbreak timing. NHs impacted early in the pandemic had greater proportions of residents recovering in isolation at the end of this study compared to those who had later outbreaks, despite both having similar proportions of residents and staff infection. This suggests more prolonged outbreaks in these facilities. Scarcity of personal protection equipment, difficulty accessing testing in a timely fashion, and knowledge of atypical symptoms and asymptomatic infection were significant factors which were more challenging earlier in the pandemic. Of concern, despite progress in these areas over the course of the pandemic, similar numbers of residents and increased proportions of staff were infected, although overall mortality rates appeared lower in NHs with later outbreaks. Interestingly, staffing numbers with “symptomatic” rather than “asymptomatic” COVID-19 correlated closely with resident infections, and may support emerging evidence that pre-symptomatic/asymptomatic carriers have lower viral loads than those with symptoms [13]. The proportions of staff infected emphasises the need for regular systematic testing in NHs.

Public NHs appeared to have been impacted disproportionately severely relative to their private counterparts, although we must be cautious interpreting this finding as only five small facilities returned outcomes (4 with outbreaks). Nonetheless, there was substantially higher infection rates and case-fatality in public facilities. This may be explained by a more complex case-mix in public NHs, where residents are frailer, with higher medical and social care needs requiring greater amounts of close-contact care [14].

This is the largest series reporting the impact of COVID-19 on NHs, residents, and staff in the literature to date. It not only includes information from a point-prevalence study, but iterative information as the pandemic unfolded. While all outbreak NHs in this series were still in an active period, few had active new cases in the interim following the end of this study period. There are some limitations to this study. Despite the size of the sample it represents 5% of all Irish NHs, and 8% of NHs with COVID-19 clusters. Information is self-reported, although senior NH medical/nursing staff were best placed to give accurate information. We did not have information on the nature, and severity of symptoms which would be of interest in light of the reports of atypical symptoms in this cohort.

## Conclusion

This study demonstrates the substantial impact of COVID-19 on NH residents and staff. Further research is required to examine factors such as the impact of staffing levels, and case-mix on outcomes, and the significance of laboratory-confirmed COVID-19 in asymptomatic individuals.

## Data Availability

Anonymous dataset available on request.

